# The perceived impact of a support programme for caregivers of children with complex neurodisability (Encompass): findings from a pilot and feasibility study

**DOI:** 10.64898/2026.02.11.26346108

**Authors:** Kirsten Prest, Kirsten Barnicot, Aleksandra J. Borek, Phillip Harniess, Cally J. Tann, Rachel Lassman, Alea Jannath, Rachel Osbourne, Keely Thomas, Melanie Whyte, Michelle Heys, Angela Harden

**Author notes:** These authors contributed equally to this work and share last authorship (Michelle Heys, Angela Harden).

## Abstract

**Purpose:** Caregivers of children with complex neurodisability frequently experience high caregiving demands, social isolation, unmet support needs, and reduced wellbeing. This paper explores caregivers’ perceptions of the impact of “Encompass”, a ten-modular, community-based group support programme for caregivers of children under five with complex neurodisability, co-facilitated by an expert parent.

**Materials and methods:** This study formed part of a pilot and feasibility study conducted in two socially disadvantaged, ethnically diverse urban areas in the United Kingdom. Outcome measures were collected pre-intervention, post-intervention and at three-month follow-up to explore caregiver wellbeing, empowerment, activation, and quality of life. Semi-structured qualitative interviews were conducted within three months of programme completion. Interview data were analysed using deductive coding informed by the “Encompass” programme theory alongside inductive analysis to explore mechanisms and unanticipated benefits.

**Results and conclusions:** Seven participating caregivers described improved wellbeing, increased confidence in caring for their child, navigating services, advocating for their family and engaging in the community. Peer support, shared learning and expert parent facilitation were key identified mechanisms of impact. Data from outcome measures showed patterns of improvement post-intervention, with less consistent effects at follow-up. Findings confirmed the key change mechanisms, informing future iterations and other caregiver group programmes.

**Trial Registration:** ClinicalTrials.gov Identifier: NCT06310681

## Introduction

Children with complex neurodisability, like cerebral palsy, require holistic, multi-disciplinary, family-centred support for optimal participation in home and community life (1). Children with complex neurodisability may find it challenging to participate in their daily activities due to difficulties with movement, posture, perception, cognition, communication, and co-morbid diagnoses or sensory needs (2). However, contemporary perspectives on childhood-onset disability recognise that interventions should move away from ‘fixing’ these impairments, and towards focusing on services that support the family goals, address parent health and wellbeing, and reduce barriers to participation (3). In essence, family-centred care is required, which involves working in partnership with caregivers to meet the tailored goals of the family (4).

Primary caregivers, most often mothers, face unique challenges as they navigate complex health, social and education systems where they often feel as though they need to ‘fight’ for services or information (5,6). These caregivers have consistently poorer outcomes relating to their physical and psychological health compared to those without children with complex neurodisability (7). The high burden of care and demands on their time contribute to poorer health outcomes (8), along with navigating grief and acceptance, sleep deprivation, financial strain and social isolation (9). In ethnically and culturally diverse urban settings characterised by high levels of socioeconomic deprivation (10), such as the boroughs in which this study took place, these challenges may be further compounded, increasing the risk of unmet needs and inequitable access to support.

Addressing some of these challenges requires strengthening caregivers’ empowerment, and activation. In this study, we conceptualise caregiver empowerment as caregivers’ perceived knowledge, confidence, and capacity to act effectively and exert influence on behalf of their child within service and community systems (11). Caregiver activation refers to caregivers’ knowledge, skills, confidence, and willingness to manage their child’s health and to work collaboratively with services in practice (12,13). Health literacy, a distinct concept but linked to activation, supports caregivers’ ability to access, understand, and use health information and services to make appropriate decisions (13,14). Within Levesque’s model of access to healthcare, health literacy, empowerment, and activation are relevant across caregivers’ abilities to perceive, seek, reach, and engage with care, and to manage the financial and practical burdens associated with accessing care (15). These concepts are especially relevant for caregivers of children with complex neurodisability, who must navigate multiple services across health, education, and social care and may have significant unmet support needs.

In our research, we use ‘complex neurodisability’ to refer to conditions characterised by motor impairment resulting from neurological causes. A key exemplar diagnosis is cerebral palsy (CP), the most common childhood-onset physical disability, affecting approximately 1 in 400 children in the UK (16). We explored the needs of those caring for children with complex neurodisability in our setting and found that caregivers require more information and knowledge about their child’s condition as well as increased support for their own wellbeing (17). Other studies have noted the importance of peer support as a protective factor for wellbeing, and parents’ desire to be connected with others in similar situations (18).

In response to these caregiver needs, we developed the “Encompass” programme, adapted from the well-established “Baby Ubuntu” programme that is widely implemented across several low- and middle-income countries (LMICs) (19). “Baby Ubuntu” is grounded in participatory, community-based approaches that are commonly valued in LMIC contexts. The adaptation therefore represents an example of a ‘decolonised healthcare innovation’ that recognises the value of knowledge generated outside high-income settings, particularly in relation to low-cost, community-based programmes (20). Although caregiver support programmes exist in the UK, relatively few have been developed in culturally and socioeconomically diverse settings. “Encompass” was co-designed with a diverse group of caregivers and piloted in urban areas characterised by ethnic, cultural and linguistic diversity and socioeconomic disparities. For this reason, we anticipate that it may be more acceptable in similarly diverse communities in the UK, and potentially in other high-income settings.

“Encompass” is a modular, community-based group programme for caregivers of children with complex neurodisability under five years old. It is co-facilitated by an expert parent with lived experience alongside a healthcare professional. It aims to improve their knowledge, skills and confidence in caring for their child, provide an opportunity for peer support networks to form, and enhance the wellbeing and overall quality of life for the caregivers, children and family.

The aim of this paper is to explore caregivers’ perceptions of the impact of the “Encompass” programme in two socially disadvantaged and diverse urban areas in the United Kingdom (UK).

## Materials and Methods

### Study Design

This paper forms part of a larger pre-post pilot and feasibility study. This article focuses on perceived impact of the programme, reporting on changes in quantitative outcomes and qualitative interviews with caregiver participants. Quantitative outcome measures were completed by caregiver participants pre-intervention, post-intervention and at three months follow-up to explore the impact of the programme on caregiver wellbeing, empowerment, activation and quality of life. Qualitative semi-structured interviews took place between November 2024 and March 2025, within three months of caregivers’ completing the “Encompass” programme. Detailed methods can be found in the study protocol (21) and the study registration on clinical trials.gov (ID: NCT06310681).

### Setting

The study took place in two London boroughs with significant ethnic, cultural and linguistic diversity. Local support for children with disabilities and their families is primarily provided by community services in the UK’s National Health Service (NHS). Although NHS care is provided free at the point of delivery, the system is currently experiencing substantial pressures, including staffing shortages and increasing service demand (22). Both boroughs experience high levels of socio-economic deprivation and health inequality (23,24), with approximately 50% of children considered to be living in poverty (10). The two study areas report some of the lowest levels of health literacy nationally (58–67%), exceeding the UK average of 41% (25). Low health literacy is a known barrier for families’ ability to access and interpret health information and is associated with poorer health outcomes (26).

### Intervention

The “Encompass” programme is an adapted version of the “Baby Ubuntu” intervention for a UK NHS setting, delivered as a 10-module, group-based programme over a six-month period (19). Sessions were co-created with caregivers of children under five with complex neurodisability. The groups are participatory in nature, meaning that caregivers actively contribute to the content and learning process, drawing on their own experiences and expertise in line with principles of adult learning theory (27). Each group is co-facilitated by a trained health professional and an “expert parent” with lived experience in caring for a child with a complex neurodisability. Modules focus on building knowledge, confidence, and peer support through shared learning, discussion, and practical activities. Children are welcomed to attend the sessions, allowing caregivers to practise and apply new skills within the group setting. Each sessions covers specific topics including child development, play, positioning, communication, everyday routines, and navigating health and education systems (figure 1). Caregiver wellbeing is addressed throughout the sessions.

**Figure 1:**
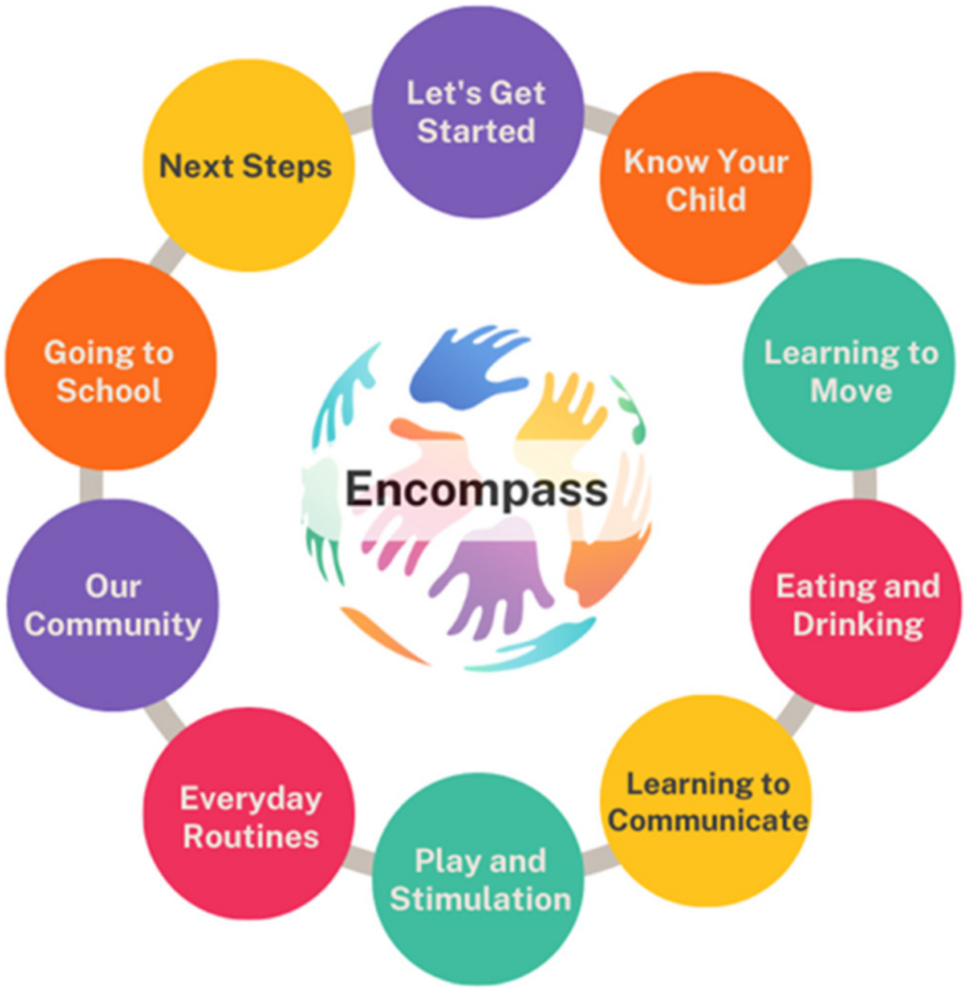
The “Encompass” modules, reproduced from Prest et al. (2025) (21)

### Involvement of those with lived experience

The recognition of expertise found in lived experience is a core value of the “Encompass” programme. It is reflected in the programme design, delivery and evaluation. Four mothers of children with complex neurodisability were not only involved in the adaptation and co-design of the programme itself, but also actively contributed at multiple stages of the research process. The aim was to ensure that the study was relevant, accessible and meaningful to those with lived experience. Activities included providing feedback on participant-facing materials, advising on recruitment approaches, and contributing to the interpretation of findings through sense-checking results and selecting quotations for publication. These activities took place in the form of group workshops either in person or online. The parent partners are co-authors of this publication in recognition of their substantial contributions to the programme and project.

### Participant selection and recruitment

Participants were caregivers of children with complex neurodisability. Full eligibility criteria for participation in the study are presented in Table 1. Eligible caregivers were approached by members of the early years therapy teams (including physiotherapists, occupational therapists, and speech and language therapists) either during clinical appointments or by telephone. They received an information sheet outlining the study and the researcher’s contact details. Caregivers who wished to take part registered their interest by emailing the researcher or scanning a QR code. Written informed consent was obtained once participants had reviewed the information and had the opportunity to ask questions.

**Table 1:** Inclusion and exclusion criteria for caregiver participants.

| Inclusion | Exclusion |
| --- | --- |
| • Primary caregiver of a child (<5 years at the time of enrolment) with a complex neurodisability*<br>• Had received a diagnosis for their child, which had been disclosed to them, even if this was a diagnosis such as SWAN (Syndrome Without A Name)<br>• Resided in the two study boroughs in the UK<br>• ≥ 18 years of age<br>• There were no exclusions based on language, as interpreting/translating services were offered | • Primary caregiver of a child with:<br>- a developmental disability where there were no functional physical impairments as part of their complex needs. For example, children diagnosed with autism spectrum disorder, attention-deficit hyperactivity disorder, and intellectual impairments were excluded unless they had a functional physical impairment with a neurological cause too<br>- a progressive neurological condition such as Duchenne’s Muscular Dystrophy<br>- a structural physical impairment not caused by a neurological event or neurological difficulties. For example, children born with a limb difference or a child with hearing loss<br>• Did not have capacity to consent |

\* *Complex neurodisability for this study is based on need over a specific diagnosis. Children should have the following: A non-progressive neurological disorder either caused by a congenital brain abnormality or an acquired long-term condition caused by a neurological event (e.g. Hypoxic-Ischemic Encephalopathy or Traumatic Brain Injury), resulting in a functional physical impairment. Additional difficulties with cognition, hearing and vision communication, emotion and behaviour can form part of the child’s clinical picture, but functional physical difficulties must be present*

### Qualitative data collection and analysis

Semi-structured interviews were conducted with all caregiver participants to explore perceptions of the impact of the “Encompass” programme. Caregivers reflected on their experiences of the “Encompass” programme, including perceptions of its content, facilitation, and group participation. The Theoretical Domains Framework (TDF) (28) informed the topic guide to ensure the interviews explored key behavioural domains. The topic guide covered changes in knowledge, confidence, and motivation, as well as environmental and social factors influencing engagement and application of skills (see appendix 1). All interviews were conducted on MS Teams or in-person by the lead researcher (KP), an occupational therapist and researcher, who had built rapport with the families over the six-month study period. Although not involved in delivering the “Encompass” sessions, KP attended weekly groups in a supportive role, assisting with logistics and acting as a consistent point of contact for caregivers. This established rapport facilitated open and honest discussion, and participants were reassured that they could be critical about the programme, and that feedback both positive and negative would be helpful at this early piloting stage. Reflexive discussions were held with members of the research team as well as the “Encompass” parent research partners.

The interviews were audio-recorded, transcribed and pseudonymised. Data analysis was managed using NVivo software (V15). Transcripts were analysed deductively by coding the data into the four categories of outcomes included in the “Encompass” programme theory logic model (Figure 2) relating to: caregiver wellbeing, confidence, advocacy, and knowledge and skills in managing their child’s health and service access. Codes that did not relate to those four predefined outcomes were analysed inductively, to see whether alternative mechanisms or outcomes of the intervention were described. Coding and theme development were led by KP, with regular discussion and review by the wider research team (AH, KB, AJB, PH), contributing to overall interpretation. Results were discussed with the “Encompass” parent partners (AJ, RO, KT, MW) for sense-checking, and to help select the most illustrative quotes for the results.

**Figure 2:**
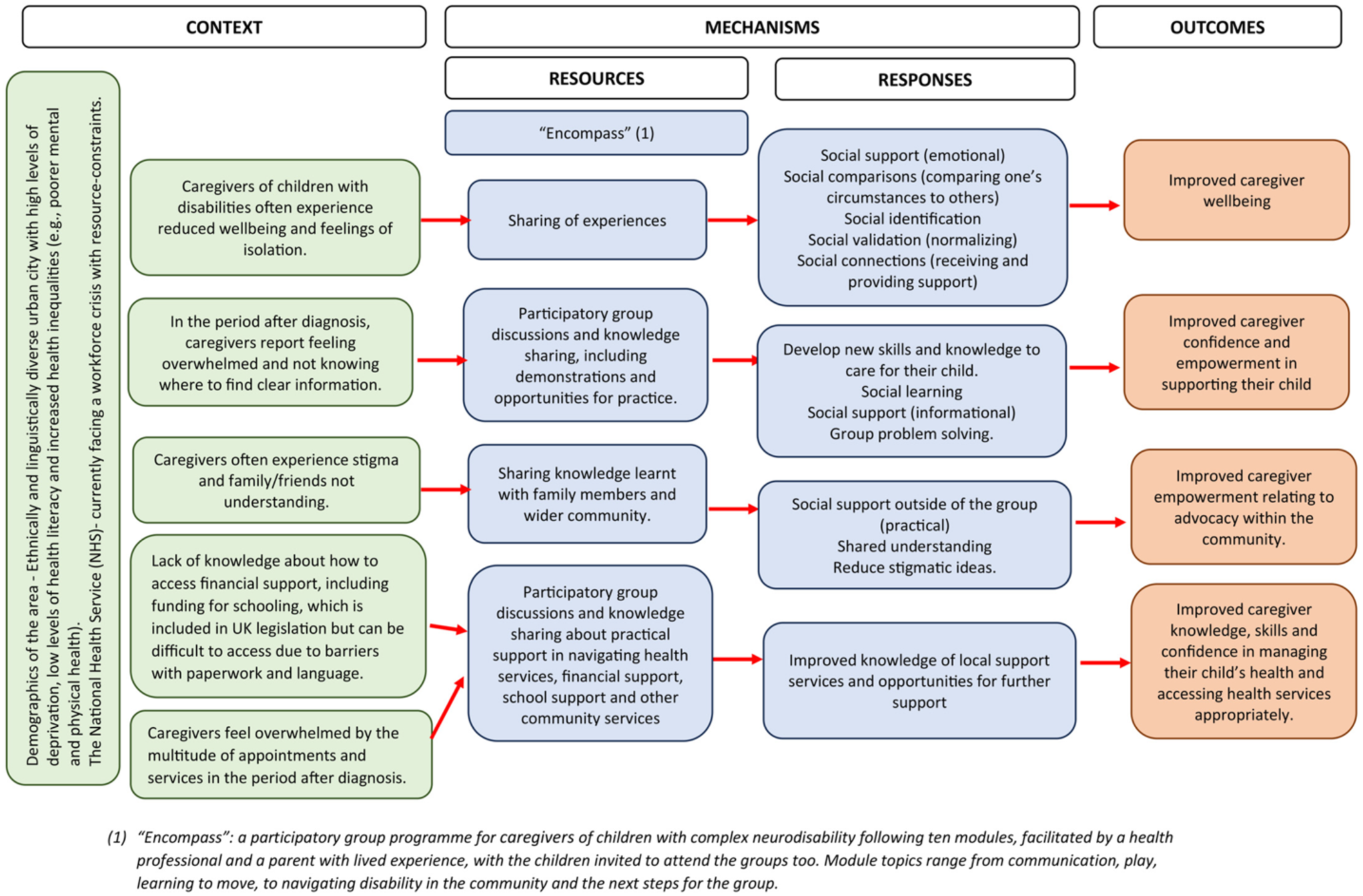
The “Encompass” programme theory logic model, reproduced from Prest et al. (2025) (19)

### Quantitative data collection and analysis

Initial indicators of programme impact were explored using quantitative individual-level outcome measures. These included the Family Empowerment Scale (FES) (29), the Parent-Patient Activation Measure (P-PAM) (30), the Warwick Edinburgh Mental Wellbeing Scale (WEM-WBS) (31), the EuroQoL 5-dimension questionnaire (EQ-5D-5L) (32) and the Power Ladder Question (PLQ) (33,34). The P-PAM reflects caregivers’ confidence, knowledge, and perseverance in managing their child’s care, which relates to to activation and health literacy. The FES assesses empowerment within the family (confidence in parenting and decision-making), within the service system (ability to navigate and influence services), and within the community (participation and perceived impact beyond the immediate family). The Power Ladder Question captures caregivers’ broader sense of power and influence over their lives. WEM-WBS provides a measure of mental wellbeing, while the EQ-5D-5L assesses overall health-related quality of life across five domains. Caregivers were also asked about their biggest issues that they face in everyday life and their main goals for attending the group (35). Further details about each outcome measure may be found in the study protocol (21).

Quantitative data collected from the individual-level outcomes were analysed using descriptive statistics and summarised using means and standard deviations. It was not appropriate to conduct statistical analyses in a feasibility study like this with such a small sample size (36).

### Integration of data

Integration of qualitative and quantitative data was guided by the “Encompass” logic model. Following separate analysis of each dataset, findings were brought together and descriptive quantitative results provided a pattern to contextualise and contrast with qualitative findings rather than to determine effectiveness. Specifically, quantitative indicators of wellbeing, empowerment, and activation were compared with qualitative accounts of change within each theme, supporting interpretation of how caregivers experienced the programme and how these experiences aligned with the predefined outcomes.

### Ethical Considerations

This study received approval from the NHS Health Research Authority (ref. 23/EM/0213). Written informed consent was obtained from all participants, who were provided information about the purpose of the study and their participation. Confidentiality and data protection procedures were followed throughout, with all data stored in accordance with the Data Protection Act 2018 and GDPR. Safeguarding procedures were in place, and facilitators were trained to support participants who might experience emotional distress during the groups.

## Results

Eight caregivers initially consented to take part in the pilot and feasibility study. One caregiver withdrew after the first session due to their child’s ill health and was unable to complete baseline data collection. No qualitative data or outcome measure data were available for this participant, and they were therefore not included in subsequent analyses.

The remaining seven caregiver participants completed the ten-modules of “Encompass” and took part in a qualitative interview within three months after the intervention had ended. Sociodemographic information for the caregivers and their children is presented in Table 2. Most caregivers had only one child, and most children were male.

**Table 2:** Demographic information of caregivers (N=7) and their children.

| **Caregivers (N=7)** | **n (%)** |
| --- | --- |
| *Age, mean (range) in years* | 30 (23-37) |
| *Female* | 7 (100) |
| *Married* | 3 (43) |
| *Ethnicity* |  |
| South Asian (including Bangladeshi and Pakistani) | 4 (57) |
| Black (including Black British and Black African British) | 3 (43) |
| *Non-UK Country of Birth* | 4 (57) |
| *No Paid Employment* | 4 (57) |
| *Receiving Disabled Living Allowance* | 4 (57) |
| *Receiving Universal Credit* | 4 (57) |
| **Children (N=6)\*** |  |
| *Age at start of programme, mean (range) in years* | 3.6 (1.1 – 5.9) |
| *Diagnosis of Cerebral Palsy* | 3 (50) |
| *Diagnosis of motor disorder secondary to genetic disorder or congenital birth condition* | 3 (50) |
| *Equipment\*\** |  |
| Static specialist seating | 5 (83) |
| Wheelchair | 4 (67) |
| Standing frame | 5 (83) |
| Leg splints | 5 (83) |
| Other (e.g. feeding tube, hand splints, bath or shower chair) | 6 (100) |
| *Under the care of occupational therapist, paediatrician, physiotherapist, speech and language therapist* | 6 (100) |

\*Missing data for one child due to caregiver not consenting to their child's medical information being collected.
\*\*More than one answer can be applied per participant.

### Changes in outcome measures

Almost all outcome measures showed improvement in caregiver activation (P-PAM), empowerment (Power Ladder Question and FES) and wellbeing (WEMWBS) immediately post-intervention (Table 3). At follow-up three months post-intervention, all scores reduced slightly. Scores on the EQ-5D-5L health index measuring quality of life reduced over time, suggesting that improvements in mental wellbeing and empowerment did not necessarily translate into better overall health status. The parent partners in the research study suggested that this pattern may reflect caregivers feeling empowered immediately after the intervention, and then potentially feeling disheartened when faced with the reality of services and support. They also reported that wellbeing is often tied to external factors, for example housing difficulties and their own physical health.

**Table 3:** Quantitative results on initial indicators of programme impact (N=7)

| Outcome measure | Pre-intervention (mean) | SD | Post-intervention (mean) | SD | Follow-up (mean) | SD |
| --- | --- | --- | --- | --- | --- | --- |
| Caregiver activation (P-PAM). Higher scores = greater activation | 70.12 (n=6<sup>a</sup>) | 16.70 | 82.42 (n=6<sup>c</sup>) | 17.15 | 78.60 (n=7) | 16.27 |
| Caregiver wellbeing (WEM-WBS). Higher scores = greater wellbeing | 49.00 (n=6<sup>a</sup>) | 12.36 | 56.33 (n=6<sup>c</sup>) | 8.64 | 53.57 (n=7) | 12.78 |
| Caregiver empowerment (FES Family Score). Higher scores = greater empowerment | 4.37 (n=5<sup>ab</sup>) | 0.77 | 4.59 (n=6<sup>c</sup>) | 0.55 | 4.32 (n=7) | 0.56 |
| Caregiver empowerment (FES Service System). Higher scores = greater empowerment | 4.43 (n=5<sup>ab</sup>) | 0.82 | 4.39 (n=6<sup>c</sup>) | 0.66 | 4.23 (n=7) | 0.58 |
| Caregiver empowerment (FES Community). Higher scores = greater empowerment | 3.56 (n=5<sup>ab</sup>) | 1.19 | 3.66 (n=6<sup>c</sup>) | 0.82 | 3.47 (n=7) | 0.99 |
| Caregiver quality of life (EQ-5D-5L). Higher scores = greater quality of life | 0.91 (n=5<sup>ab</sup>) | 0.07 | 0.78 (n=6<sup>c</sup>) | 0.20 | 0.72 (n=7) | 0.29 |
| Caregiver empowerment (Power Ladder Question). Higher scores = greater empowerment | 5.50 (n=6<sup>a</sup>) | 2.95 | 7.17 (n=6<sup>c</sup>) | 1.33 | 6.71 (n=7) | 1.60 |

*Missing data: a= outcome measure questionnaires lost in the post, b= caregiver unable to complete questionnaires due to caring for their child during the group session, and c= caregiver did not attend final group session.*
*P-PAM= Parent-Patient Activation Measure, WEM-WBS= Warwick Edinburgh Mental Wellbeing Scale, FES= Family Empowerment Scale, EQ-5D-5L= EuroQoL 5-dimension questionnaire*

Five out of the seven participants provided information relating to their primary daily challenges and goals for attending the group (Table 4). Most participants reported achieving (n=4) or partially achieving (n=1) their goals. Participant goals primarily related to the outcomes of ‘improved confidence in caring for their child’ and ‘expanded social support networks’.

**Table 4:** Summary of caregiver daily challenges, goals, and perceived goal achievement (N=5)

| Questions asked to the participant | Summary of participant responses (N=5)* |
| --- | --- |
| Biggest issues in daily life that you need support with | High caring demands, limited personal time, difficulties balancing exercises and routines, challenges with their child’s communication, feeding or mobility needs, disturbed sleep, housing issues, and working with carers. |
| Main goals for attending the group | Increase their knowledge and confidence, learn from others’ experiences, share their own experiences, build connections, and support their child’s communication. |
| Do you feel that you have achieved your goals? | Four caregivers reported that they had achieved their goal. One caregiver described partially achieving their goal, as ongoing challenges in finding time for themselves continued. |

*\* Data were unavailable for two participants due to loss of questionnaires in the post and because one caregiver was unable to complete the questionnaires while caring for their child during the group session.*

### Integration of findings

Table 5 summarises the five outcomes and sub-categories that were identified in the qualitative analysis, showing how each relates to the original “Encompass” programme theory. The table indicates whether sub-categories represent (i) mechanisms of change (processes explaining how change occurred) or (ii) expressions of outcomes (specific, observable changes, benefits, or results of the programme). Sub-categories are also labelled as :

- *Aligned* with existing programme theory, where findings (mechanisms or outcomes) were consistent with elements already present in the original logic model that represented the programme theory
- *Adds detail* to existing programme theory, where findings elaborated on existing mechanisms or outcomes
- *Extends* the original programme theory, where findings introduced new mechanisms or outcomes not included in the original model.

**Table 5:** Overview of outcomes, sub-categories, and their relationship to the programme theory.

| **Outcomes** | **Sub-Categories** | **Mechanism/**<br>**expression of**<br>**outcome** | **Relation to**<br>**programme theory** |
| --- | --- | --- | --- |
| Improved caregiver wellbeing | Reduced isolation and a sense of belonging | Mechanism | Aligned |
|  | Opportunities for emotional expression and support | Mechanism | Aligned |
|  | Increased hope and reduced fear of the future | Mechanism | Adds detail |
|  | Feeling seen and cared for | Mechanism | Adds detail |
|  | Improved self-care | Expression of outcome | Adds detail |
| Improved confidence in caring for their child | Gaining practical knowledge and strategies | Mechanism | Aligned |
|  | Feeling reassured and validated in existing practices | Mechanism | Adds detail |
|  | Increased confidence in communicating with their child | Expression of outcome and mechanism | Aligned |
| Improved confidence navigating health services | Improved understanding of specific processes, support and terminology | Mechanism | Aligned |
|  | Increased confidence to engage with and negotiate services | Expression of outcome and mechanism | Aligned |
| Improved empowerment, advocacy, and participation in the community | Increased empowerment and confidence to advocate | Expression of outcome and mechanism | Aligned |
|  | Increased participation and engagement in community life | Expression of outcome | Extends |
| Expanded social support networks (new outcome) | Building connections with caregiver who share similar experiences | Mechanism of new outcome | Extends |
|  | Continued connection and expanding networks beyond the group | Expression of new outcome | Extends |

#### Outcome 1: Improved caregiver wellbeing

Caregivers described emotional and psychological benefits after attending the group, consistent with the programme theory’s outcome of improved wellbeing. The sub-categories within this outcome illustrate how the group fostered wellbeing (mechanisms such as reduced isolation and opportunities for emotional expression) and what this improved wellbeing looked like in practice for caregivers (expression of the outcome such as an increase in self-care activities). The group wellbeing scores (WEMWBS) showed an increase immediately after the intervention, followed by a slight decline at follow-up.

##### Reduced isolation and a sense of belonging (mechanism)

Across the interviews, caregivers consistently described entering the group feeling isolated or unsupported, and the subsequent relief that they experienced when meeting others with similar experiences. This created a sense of connection and belonging and reduced feelings of being alone in their difficulties.

> *“The [most significant] change is that I realise that I am not alone… I’m not the only parent who have a child with disability. I need to have more effort to wake up and get out.” (C8)*

##### Opportunities for emotional expression and support (mechanism)

Feeling a connection to others enabled caregivers to speak openly about internalised emotions. The practical, reflective and visual activities helped caregivers to articulate emotions that they had struggled to express before. Hearing others’ experiences and sharing their own helped to normalise difficult feelings.

> *“The string one [game where you used a piece of string to visually represent your journey of having a child with a disability] was great in terms of emotional expression. It really demonstrated my feelings as a parent.” (C3)*

> *“I’m a kind of person who doesn’t share or speak, I keep everything for myself so it was a bit difficult…but afterwards I feel good, I feel very good to share and to, like empty my [gesturing to chest or heart] heart. Yes, because I was keeping in everything, no friend to share. So yeah, it was very helpful to share my situation with the group.” (C8)*

##### Feeling seen and cared for (mechanism)

Caregivers described important moments in the group where they felt noticed and cared for. Having a warm and welcoming atmosphere as well as food and refreshments signalled to them that their needs had been considered, and that they were valued.

> *“That was nice because no one sees us, we just do it for ourselves and the kids. So to walk in and have, like, a table full of food it’s like, oh my God, somebody actually has seen us.” (C6)*

##### Increased hope and reduced fear of the future (outcome expression)

Many caregivers described feeling more reassured about their child’s future after meeting the expert parent facilitators and their older children. Although emotionally overwhelming at first, seeing an example of a young person living well into adolescence helped parents imagine longer term possibilities. Being able to envision a future that includes hope was a contributor to improved wellbeing.

> *“[The expert parent facilitator] has got more experience than most of us, and it’s so nice to hear that she’s over the hard and it really inspired me and gave me hope that I’m going to be over the [hard] and that it will just be so normal to have the extra bit that that we need to do.” (C5)*

> *“When I first saw [parent facilitator’s child], I was about to cry because like I said I see my child in them when they become more big.. seeing them in the chair and not able to do things like other children it was hard.. but also seeing that someone with cerebral palsy can live long, you know it help a lot.” (C8)*

##### Improved self-care (outcome expression)

Participation in the group prompted some caregivers to prioritise their own needs for the first time in a long while. It helped caregivers gain the confidence to engage in self-care activities, have patience with themselves and make decisions that benefitted their own wellbeing.

> *“I’ve just started doing more things for myself… going to the gym… spending more time on myself.” (C1)*

> *“I think I’ve just given myself a little bit more grace…as much as I’m patient with [my child], I must also be patient with myself.” (C5)*

##### Ongoing challenges despite participation

While many caregivers experienced improvements in wellbeing, one participant reported persistent difficulty due to longstanding depression, anxiety, and chronic pain. Although she valued the group and felt happier during the sessions, these broader challenges continued to shape her daily life and limited the extent of her overall improvement, illustrating that gains in wellbeing were not experienced by all. This is consistent with the decline observed in EQ-5D-5L scores, which may partly reflect the impact of ongoing physical health problems for some caregivers.

> *“I’ve been depressed… I’ve had anxiety… I didn’t want to go out. My pain is in my back, it’s very cramped and I can’t cope with it.” (C3)*

#### Outcome 2: Improved confidence in caring for their child

This theme reflects both increases in caregivers’ confidence and the processes through which this confidence was built, consistent with the logic model’s outcome of improved skills, confidence and knowledge in caring for their child. Caregivers described learning practical strategies, gaining reassurance about existing skills, and improving their confidence in communicating with their child.

##### Gaining practical knowledge and strategies (mechanism)

Caregivers described becoming more confident after learning concrete, practical strategies for supporting their child’s communication, feeding, positioning, and daily routines. They also described learning useful information relating to schooling. Demonstrations and guidance from the facilitators and others helped them understand how best to take care of their child. Caregivers reported that “Encompass” differed from other workshops or courses that they had been a part of, due to the fact that a parent with lived experience was a facilitator and could understand how they might be feeling.

> *“I felt really like empowered and knowledgeable, as if I was better equipped to parent [my child] because I think with every session I left with like learning something new or even if there’s those sheets where you’d print and give them out, I was like, I can’t wait to go home and tell my husband about today’s session and go and tell him what we learned.” (C2)*

> *“I remember how [the facilitators showed us]… how to hold the children…and how to help them to sit correctly and how to position them.…how to lay them…very helpful and the feeding as well, how to use the cup” (C8)*

##### Feeling reassured and validated in existing practices (mechanism)

Several participants explained that the sessions confirmed they were already doing many things well. Having their existing approaches to caring for their child acknowledged and normalised by others in the group reinforced their sense of confidence and competence. For example, caregivers realised their confidence and capability in positioning their child, and supporting them with their movement.

> *“I was doing everything that was highlighted in that module… I thought oh wow, I’m actually doing things right.” (C1)*

> *“It also highlighted to me that my own personal experience and all the learning that I do at home, is like self-sufficient. I gave myself a pat on the back when I left, I was thinking like I really do the work for my child, I really dig deep and do the homework.” (C6)*

##### Increased confidence in communicating with their child (mechanism and outcome expression)

Learning new ways to interpret and respond to their child’s communication, such as using gestures and visual supports, helped caregivers feel more connected to their child’s needs. These strategies strengthened their confidence in understanding and supporting their child.

> *“The module that stands out for me is learning to communicate… I didn’t really think about it from [my child]’s point of view… So that kind of really opened my eyes. That was the one that was kind of pivotal for me, like what I’m going to do different going forward.” (C2)*

#### Outcome 3: Improved confidence in navigating services

Caregivers described increased confidence in navigating complex health systems, consistent with the logic model’s proposition that the group would improve their confidence to manage their child’s many health services. Two mechanisms supported this change, including improved understanding of services and systems, and feeling more confident to engage with and negotiate with services. These interview findings correspond with small increases on the Family Empowerment Scale (FES) service-system scores immediately post-intervention, although these were not sustained at follow-up. Increases were also seen on the Parent Patient Activation Measure (P-PAM), which measures caregivers’ confidence and engagement in managing their child’s health, providing tentative quantitative alignment with these qualitative findings.

##### Improved understanding on processes, support and terminology (mechanism)

Some caregivers described gaining a clearer understanding of particular aspects related to their child’s care, such as elements of the Education Health and Care Plans (EHCP) process, where to find further support, or medical terms used by professionals to describe their child’s condition.

> *“There was the our community [module], which I found really helpful because that just highlighted kind of all the other help that you could get, which I don’t think I would have thought of or knew about. So I really learnt a lot there.” (C2)*

> *“These medical terms, we didn’t even hear about this type of disorder before in our life so now I know…about [child’s name] and their medical terms.” (C7)*

##### Increased confidence to engage with and negotiate services (expression of outcome and mechanism of change)

Hearing how other parents advocated for their children helped participants recognise that they did not need to *“sit and wait”* (C2), but could ask questions, negotiate alternatives, and ask for support when needed. Several described feeling more able to request different appointment times or formats, reflecting an increased empowerment in advocating for their child’s needs and their own.

> *“And ask about whether we can do teams appointments, [that] was a good one… now like I have the confidence to be like actually if I did have a health visitor appointment, for example, I know I can get another one that week.” (C2)*

#### Outcome 4: Improved empowerment, advocacy, and participation within the community

This theme reflects both the empowerment processes expected in the original logic model and an extension of the model to include community participation as part of the outcome. Participation for the caregiver and child appeared to come from increased confidence and a stronger sense of agency. Quantitative scores on the FES community subscale and the Power Ladder question showed slight post-intervention increases, consistent with caregivers’ descriptions of improved confidence and a stronger sense of influence within their communities.

##### Increased empowerment and confidence to advocate (mechanism and outcome expression)

Caregivers described developing a stronger sense of empowerment, influenced by others in the group and by reflective activities in the sessions. Many reported feeling more confident in speaking up, sharing their stories, challenging misconceptions, and supporting others. For some, this extended into taking on new roles within their child’s school or wider parent networks and recognising themselves as capable advocates or community leaders. Several participants also noted that they engaged more with family, friends, neighbours, and other parents at their children’s schools through sharing information and encouraging a greater understanding of disability.

> *“I think I’ve become a lot more confident since joining the group because it rubs off on you how the others fight for their kids and you know…you don’t have to just sit and wait, you can actually ask and fight for it, I haven’t really had to do all of that, but hearing how the others have done it gives you a bit of that firepower.” (C2)*

> *“Yes, it was very helpful because before, I didn’t know that I was able to write to a MP, yes, I find out that in the groups that you can write to the MPs for help.” (C8)*

##### Increased participation and engagement in community life (new expression of outcome)

Alongside greater advocacy, many caregivers described becoming more willing and able to engage in community settings. This included taking their child out more often, reconnecting with local groups, sharing resources with neighbours, and in some cases planning future education or employment. These shifts reflected a move from isolation to community participation, which links to the improved wellbeing outcome too.

> *“I want to go back to college… improve my English and maybe do health and social care.” (C8)*

> *“It really clarified that I have the confidence to be a spokesperson and speak up and I can actually lead if I wanted to. From the group I now have gone on to… work in my child’s school… it really opened doors.” (C6)*

#### Outcome 5: Expanded social support networks

Although social support can be understood as a mechanism underlying several of the outcomes above, caregivers described such significant changes in their social support networks, that it was considered as a distinct outcome. Two subthemes capture the mechanisms as to how these peer support networks were developed and sustained beyond the end of the programme.

##### Building connections with caregivers who share similar experiences (mechanism)

Caregivers consistently described forming supportive relationships with other parents in the group. Many reported that it felt easier to open up to people who “understood” because their children had similar disabilities, reducing feelings of isolation and creating a sense of belonging. Connecting in informal moments, during the breaks in the group, through shared stories within the activities, or by meeting one another’s children, helped strengthen these connections. For several participants, this was their first experience of having peers they could relate to, which brought a sense of comfort, reassurance and belonging.

> *“I felt like I had this extra support system that I didn’t have before because most of the parents were further on in their journey than I was. So it was really nice to see how they’re doing and the advice they’d give. It’s not something I would have found anywhere else. And it’s really hard like to even find SEND mums, let alone ones with neuro disabilities” (C2)*

##### Continued connection and expanding networks beyond the group (new outcome expression)

Many caregivers maintained contact after the programme ended, through WhatsApp groups, informal meet-ups, and ongoing conversations, and some extended these networks into their wider communities. Several participants described actively facilitating new connections for others, indicating a shift from receiving support to generating it.

> *“We’re still on the WhatsApp group. So that’s really nice to hear from them and I think I asked them a question as well and they answered it, so I was like still seeking help, so that’s been really good.” (C2)*

## Discussion

These findings demonstrate the positive impacts that a low-cost community-based caregiver support programme can have at an individual and family level as well as in their wider communities. According to caregivers, their wellbeing improved through reduced isolation, feeling seen and being able to express their emotions, having hope for the future and caring for themselves more. Confidence in caring for their child and navigating services increased along with increased empowerment, advocacy and community participation. A new outcome was identified for the programme theory: the expanded and sustained social support networks for caregivers.

It is well documented in literature that peer support amongst caregivers of children with disabilities can reduce feelings of isolation, increase the sense of belonging and feeling seen and heard (37–40). Group-based interventions may generate change through processes such as sharing of experiences, social support (informational, emotional and practical), social validation or normalisation, and social comparisons, which reduce isolation and build supportive networks (41). It was clear in this study that caregivers experienced these mechanisms which had an impact on their wellbeing. As a result of this programme, peer support went beyond the group, and a community was built that continued to meet afterwards and remain in contact. Higher levels of perceived social support are associated with better caregiver wellbeing, including lower parenting stress, reduced depressive symptoms, and lower psychological distress among caregivers of children with disabilities (42,43).

A significant finding from this study was how the expert parent represented a powerful image of the caregivers’ potential future, which both encouraged participants to emulate them and helped reduce uncertainty and foster hope. Other models using these approaches have reported similar mechanisms, with parents describing experienced peers as credible role models who normalise difficult experiences, reduce uncertainty about the future, and offer reassurance through having been through similar situations (39,44,45).

While caregivers reported meaningful improvements in wellbeing through interviews and outcome measures, quality of life scores did not increase. Our data indicates that caregivers’ experience of chronic pain was one explanation of this finding. Physical health difficulties and persistent mental health difficulties, such as depression, may therefore blunt the impact of the intervention. Furthermore, the intervention is not designed to address these and access to specialist services are required. It does however raise the question of how best to quantitatively measure the impact of interventions such as “Encompass”. This issue has been explored by McGlinchey et al. (2025) (40), who found that commonly used wellbeing measures may fail to capture outcomes that parents and caregivers themselves consider most meaningful, particularly when measures prioritise positive mental wellbeing, and overlook ongoing stress, physical health, and contextual burdens.

The caregivers in this study described learning practical strategies and knowledge from each other and having their existing caregiving skills validated as empowering aspects of the programme. These mechanisms are consistent with social cognitive theory, which suggests that self-efficacy, or confidence in one’s ability to manage challenges, develops through shared learning and observing others in similar situations (46). In this study, increased confidence and empowerment appeared to be associated with greater participation in community life for some families, extending the original programme theory. Families of children with complex neurodisability often face barriers to participation that are not only structural but may also be shaped by internalised stigma, fear and anxiety (47). Participants described how seeing other caregivers confidently engaging in community activities offered reassurance and a sense of possibility. Similar participation-related outcomes have been reported in evaluations of parent-led support groups and peer support programmes (48,49).

The perceived impacts reported by caregivers in this study are consistent with mechanisms described across other interventions originating from the “Ubuntu” programme model. For example, evaluations of “Getting to Know Cerebral Palsy” have reported improvements in caregivers’ knowledge and confidence alongside improved family functioning and parental quality of life. Qualitative work highlighted hope and the sense of not feeling alone as key mechanisms of improved wellbeing (50,51). Similarly, “Juntos” has emphasised the value of peer facilitators, with feasibility studies describing expert mothers as credible sources of encouragement and shared learning, and reporting improved caregiver knowledge and confidence following programme participation (52,53). Mixed methods evaluations within this programme model also highlight challenges in consistently demonstrating change quantitatively. Earlier pilot work linked to the “Baby Ubuntu” programme reported approximately a 25% improvement in family quality of life scores (54), whereas the later mixed methods randomised feasibility trial did not find conclusive change on the same measure despite strong qualitative evidence of impact (55).

### Implications for programme development and clinical practice

This paper provides important results for the further development and evaluation of the “Encompass” programme and for the design of caregiver support programmes more broadly. It highlights the significance of the expert parents, including the value of them bringing their children to the group so that caregivers of younger children can glimpse what the future may hold. It also suggests that a nurturing environment and hospitality can foster a sense of safety and belonging, where caregivers feel seen and cared for, and may represent an important mechanism of the programme. This is likely to be relevant for other group-based caregiver interventions too. The programme theory depicted through the logic model will undergo further extension and exploration with an advisory group to incorporate the results from this study. More generally, increased community participation and social support as outcomes should be explored further across caregiver programmes, along with careful consideration of how best to measure these constructs.

For clinical services, detailed findings on the feasibility and acceptability of the programme delivery and evaluation within NHS community settings will be published elsewhere. This paper demonstrates the value of group-based support programmes for caregivers of children with complex neurodisability in ethnically and culturally diverse urban areas with high levels of social deprivation. Caregivers not only learned valuable therapeutic skills relating to positioning, feeding, play and communication but they better understood services, systems and how to advocate for their children. Given that clinicians often go beyond formal clinical roles to support families to understand services, navigate systems and access support, interventions such as “Encompass” may offer a complementary way of supporting this work by building caregiver confidence and understanding.

### Strengths and Limitations

This study is strengthened by its mixed methods design, which enabled comparison across qualitative and quantitative data. The involvement of caregivers with lived experience throughout the project was another strength which ensured the research was relevant. Findings were broadly consistent with the programme theory and provide the basis for refining the mechanisms of change. As a pilot and feasibility study, the sample size was small, and the study was not designed or powered to detect statistical effects. The lack of a control group further prohibits drawing conclusions from the quantitative data. The researcher’s dual role in organising the programme (although not delivering it) and data collection may have influenced what participants shared in interviews despite best efforts to minimise this through reflexive practice and by emphasising to participants that honest and critical feedback was welcomed and valuable.

### Conclusion

This study highlights the meaningful impacts that a community-based support programme can have for caregivers of children with complex neurodisability. Caregivers described improvements in wellbeing, reduced isolation, increased confidence and empowerment, and strengthened peer relationships that extended beyond the group. The findings underscore the importance of expert parents, peer support and a safe, welcoming environment as key mechanisms of impact, and suggest that community participation and sustained social support may be important outcomes for future evaluation. The mixed methods findings also raise important questions about how best to measure impact in this population, particularly where commonly used quantitative tools may fail to capture changes that caregivers value. Together, these findings contribute to the emerging evidence base for group-based, caregiver support programmes and provide focused evidence for the continued development and refinement of “Encompass” for caregivers of children with complex neurodisability.

## Supporting information

Topic guide for caregiver participants

## Author contributions

KP, AH, and MH conceptualised the paper and were involved in funding acquisition. The methodology was developed by KP, KB, AJB, PH, CJT, MH and AH. Data were collected by KP and analysed by KP, KB, AJB, PH and AH. The project administration was undertaken by KP, with AH, MH and KB supervising. MW, AJ, RO and KT provided expertise from lived experience. KP, KB, AJB, PH and AH drafted the original manuscript, and all authors contributed to reviewing and editing it.

## Acknowledgements

The first author (KP) was funded by the HARP PhD Programme to conduct this research. We would like to thank members of the Encompass advisory group for their ongoing support and expertise throughout the research project including Frances Badenhorst, Dr Aleksandra Borek, Dr Phill Harniess, Alea Jannath, Rachel Lassman, Prof Christopher Morris, Rachel Osbourne, Prof Cally J Tann, Assoc Prof Tracey Smythe, Keely Thomas, Melanie Whyte and Dr Emma Wilson. Finally, thank you to the mothers who generously gave their time to participate in this study.

## Data availability statement

The data that support the findings of this study are available from the corresponding author, upon reasonable request. The data are not publicly available due to privacy or ethical restrictions.

