## Supplementary material for "The perceived impact of a support programme for caregivers of children with complex neurodisability (Encompass): findings from a pilot and feasibility study": Topic guide for caregiver participants

### Appendix 1: Topic Guide for caregiver participants

Guide for interviewer:

- Bullet pointed items are key questions to ask – these are open questions wherever possible.
  - *They are followed by suggested probes to use to obtain more detail from the participant – the interviewer may not need to use these probes if the participant has already spontaneously discussed these particular aspects of their experience*
- The questions become more specific as each section progresses - the interviewer may not need to ask later bulleted questions in each section if the participant has already covered them earlier in the conversation

**General experience of the Encompass groups**

- What was your experience of attending the Encompass groups?
  - What did you like/dislike about attending the groups?
  - Were there any barriers to attending the groups?
- How did you find the different module topics?
  - Which ones were more relevant for you and which ones were less so?
- When and how did you first find out about the groups?
  - What made you decide to join?
  - Did you have any concerns or apprehensions before joining? How did those change/remain as the groups progressed?
  - Do you have any ideas about how to make the groups more inclusive? To reach a wider group of people?
- How did you find the facilitation?
  - What was your experience of having two facilitators with different skill sets and backgrounds?
  - What did the facilitators do that was helpful?
- How did you feel going to the groups?
  - What were your thoughts and feelings before you went each session? Did this change at the end of a session?
  - What motivated you to go?
  - How did you feel during the group sessions?
  - Were there any negative aspects to the groups?
  - Why do you think some parents/carers may not attend these groups?

**Capability**

*TDF – knowledge, skills, memory, attention and decision processes*

- Have you seen a change in yourself since attending the groups?
  - For example, how you understand things, or changes in routine etc.
- What were the most important things you learnt from the groups?
  - Are there any new skills that you have gained?
  - Can you provide examples of how the groups may have influenced your understanding of your child's disability and their specific needs?
  - Are there any gaps in your knowledge? How could the group have addressed these?
- How easy or difficult have you found carrying out what you learnt from the groups?

**Opportunity (environmental factors)**

*TDF – environmental context and resources, social influences*

- What helped or hindered you from attending the groups?
  - What barriers might other parents/carers have faced in attending the groups?
  - What could we do to address these barriers?
- How did you find sharing what your learned with your family and friends?
  - What was easier to share and what was more difficult?
  - How did they respond?
  - Do you feel that there are in changes as to how your family and friends view your child?
- Were there any barriers to carrying out the skills that you learned during the groups?
  - What made it easier or difficult to practise the skills that were learned in the group?
- How did you find connecting with other parents in the group?
  - Did you share similar experiences with other parents/carers?
  - Did you keep in contact between groups?
  - What helped you to connect? What made it difficult? (e.g. language)

**Motivation (attitudes and beliefs)**

*TDF – Beliefs about capabilities (confidence) and consequences, Intentions (I plan to), Goals (I want to), emotions*

- Have you noticed any changes in your confidence as a parent/carer after the groups?
  - If so, what might these be?
  - What might make you feel more confident?
- What do you think will happen if you continue to carry out the skills learnt during the groups?
  - What are your continued goals for your child?
  - What might the overall impact on your child be?
- How did the groups make you feel?
  - What parts of the groups might have helped or hindered wellbeing?
  - How do your feelings at the time affect whether you are able to carry out the skills learnt during the groups?
